# Long-lasting effects of insufficient sleep on neurocognitive development in early adolescence

**DOI:** 10.1101/2022.01.10.22269013

**Authors:** Fan Nils Yang, Weizhen Xie, Ze Wang

## Abstract

**Importance:** Adolescents nowadays often get insufficient sleep. Yet, the long-term adverse effects of sleep loss on developing brain and behavior remains unknown.

**Objective:** To determine whether insufficient sleep leads to long-lasting impacts on mental health, cognition, and brain development in adolescents across two years.

**Design:** This longitudinal study utilized a public dataset, the Adolescent Brain Cognitive Development (ABCD) study, which is an ongoing study starting from 2016.

**Setting:** Data were collected from 21 research sites in the U.S.

**Participants:** 11,875 9-10-year-olds were recruited using stratified sampling in order to reflect the diversity of the U.S. population.

**Intervention:** Individuals with sufficient versus insufficient sleep (< 9 hours per day for adolescents) were compared after controlling for age (months), sex, race, puberty status, and other 7 covariates based on propensity score matching.

**Main Outcomes and Measures:** Behavior problems, cognition, mental health assessments, resting-state functional connectivity, gray matter volume, cortical area, cortical thickness, and structural connectivity (Fractional anisotropy) were collected and preprocessed by the ABCD study. Independent-sample t-tests and meditation analysis were performed to investigate the effects of insufficient sleep.

**Results:** 3021 matched pairs (50.7% male) were identified based on baseline assessment, with mean (SD) age of 119.5 (7.5) months. In baseline, sufficient sleep is associated less behavioral problems on 18 of 20 assessments, e.g. depress (95% CI of mean difference: -0.28 to -0.47, false discovery rate (FDR)-corrected *p*□<□.001, Cohen’s d = -0.20), better cognitive performance on 7 of 10 assessments, such as crystal cognition (95% CI: 0.81 to 1.50, FDR-corrected *p*□<□.001, Cohen’s d = 0.17), better functional connection between cortical regions and basal ganglia (all FDR-corrected *p*□<□.05, Cohen’s d >0.15), and large structure in ACC and temporal pole (all FDR-corrected *p*□<□.05, Cohen’s d >0.09). Similar patterns of effect of sufficient sleep were found in FL2 (749 pairs remained) e.g. Cohen’s d of function connectivity at baseline was correlated with Cohen’s d of that at FL2 (*r* = 0.54, 95% CI: 0.45 to 0.61, *p* < 1e-10). Mediation and longitudinal mediation analysis revealed that identified brain measures (e.g. gray matter volume of left temporal pole) at baseline mediated the effect of sufficient sleep on behavioral assessments (e.g. crystal cognition) at baseline and at FL2 (95% CI did not encompass 0, *p* < 0.05 on 100,000 random-generated bootstrapped samples).

**Conclusions and Relevance:** These results provide strong population-level evidence for the long-lasting detrimental effects of insufficient sleep on mental health, cognition, and brain function and structure in adolescents. The current study identified potential neural mechanisms of adverse effect of insufficient sleep in adolescents, which might provide a theoretical grounding for sleep intervention programs to improve the long-term developmental outcomes in adolescents.

**Key points:** *Question:* What are the long-lasting effects of insufficient sleep on neurocognitive development in early adolescents? Findings
In this propensity score matching study that included 11,875 9-10-year-old participants, we identified long-lasting adverse effects of insufficient sleep on depression, thought problems, crystal cognition, neural network connectivity, cortical areas, and gray matter volume across 2 years. Meaning
This study elucidated the neural mechanisms underlying the long-term detriments of insufficient sleep on mental health and cognition, suggesting potential intervention targets to improve sleep-related outcomes.

## Introduction

Adolescents of recent generations frequently report a shorter sleep duration and poorer sleep quality (Gariepy et al., 2020; Matricciani et al., 2012; Williams et al., 2013). As the brain maturation process is vulnerable to sleep loss (Born and Wilhelm, 2012; Dumontheil, 2016; Tononi and Cirelli, 2014, 2006), adolescents with insufficient sleep often show compromised neurocognitive functions, manifested as poorer academic performance and reduced social-emotional skills (Astill et al., 2012; Cassoff et al., 2014; de Bruin et al., 2017; Owens, 2014). However, the neural mechanisms underlying these adverse effects of sleep loss on adolescent development remain poorly understood (Dutil et al., 2018). Several covariates, such as socioeconomic status (Felden et al., 2015), sex (Lee et al., 1999), and pubertal change (Sadeh et al., 2009), can substantially influence adolescents’ sleep patterns and their neurocognitive functions, making it unclear how sleep loss directly impacts neurocognitive development. Furthermore, based on the previous cross-sectional evidence (de Bruin et al., 2017), it is unknown whether insufficient sleep at a younger age has a temporary or more long-lasting impact on adolescent neurocognitive functions. Clarifying these issues is critical not only for understanding the neurocognitive vulnerability and resilience under sleep loss in the developing brain (Krause et al., 2017), but also for theoretically grounding sleep intervention programs to improve the long-term developmental outcomes in adolescents (Wahlstrom and Owens, 2017).

To address these issues, the current study leverages data from a large-sample longitudinal study, namely the Adolescent Brain Cognitive Development (ABCD) study (Casey et al., 2018), to investigate the effects of sleep duration on neurocognitive development over 11,000 pre-adolescents or adolescents at baseline (9-10 years old), 1-year, and 2-year longitudinal follow-ups. We first identified individuals with sufficient versus insufficient sleep (< 9 hours per day for adolescents (Paruthi et al., 2016) based on the baseline measure that captures a child’s sleep duration in the past 6 months at initial testing. We then matched these two groups of individuals on various key covariates, such as age, sex, race, socioeconomic status, and puberty, based on propensity score matching procedures (Ho et al., 2011; Stuart, 2010). By minimizing potential confounds from these covariates, this approach allows us to estimate the causal impacts of baseline sleep loss on neurocognitive developmental outcomes indexed by behavioral assessments of cognitive and affective functions, rest-state functional connectivity (rs-FC), and brain structure measures at baseline, 1-year, and 2-year follow-ups. Furthermore, to evaluate the biomarkers of insufficient sleep on compromised adolescent neurocognitive development, we investigated how the changes in rs-FC and brain structure measures mediate the relationship between sleep loss and reduced cognitive and affective functions.

## Methods

### Data source

The data used in the current study is from the ABCD data release 3.0 (https://abcdstudy.org), which is composed of behavioral and neuroimaging data from 9- to 10-year-old children recruited across 21 sites in the US at baseline assessment (*n* = 11,878), 1-year follow-up (FL1, *n* = 11,235), and 2-year follow-up (FL2, *n* = 6,571). Detailed protocols and designs have been previously described (Casey et al., 2018). Informed consent from the primary caregiver and assent from children were obtained before the study. This project has been approved by the Institutional Review Boards of local study sites.

### Measurements

Of primary interest, we extracted parent-reported sleep duration of the children as the independent variable. According to the American Academy of Sleep Medicine, sufficient sleep for children aged 9-12 years is 9-12 hours per day (Paruthi et al., 2016). Hence, in the current study, children with less than 9-hour sleep per day were considered with insufficient sleep (IS group), whereas children with no less than 9-hour sleep per day were considered with sufficient sleep (SS group). Dependent variables included assessments of the children’s problem behaviors, mental health, neurocognition, and brain measurements of network connectivity (a measure derived from rs-FC), cortical area, gray matter volume, cortical thickness, and structural connectivity (fractional anisotropy, FA), afforded by the ABCD dataset. Participants’ age (in month), sex at birth, race,body mass index (BMI), puberty status (1-4, assessed by *ABCD Youth Pubertal Development Scale and Menstrual Cycle Survey History*), highest educational achievement of the primary caregiver, household income, and location (study site) were considered as covariates (see further details in *Data Analysis*).

#### Sleep duration

The parent-reported *Sleep Disturbance Scale for Children* was administered at baseline, FL1, and FL2 to assess a child’s sleep quality (Bruni et al., 1996). We used the data from the question, “how many hours of sleep does your child get on most nights in the past six months?” to obtain a measure of the children’s sleep duration. Based on prior work (Paruthi et al., 2016), we re-coded this sleep duration measure to 1 for >= 9 hours sleep per day and to 0 for < 9 hours sleep per day.

#### Problem behaviors

The parent-reported Child Behavior Checklist (*cbcl*) was used at baseline, FL1, and FL2 to capture a child’s behaviors in emotional, social, and behavioral domains (Achenbach and Rescorla, 2001). Data from 20 available assessments were extracted from this checklist (see Supplementary Table S1 for the variable table), capturing the following syndrome or issues: anxious/depressed, withdrawn/depressed, somatic complaints, social problems, thought problems, attention problems, rule-breaking behavior, aggressive behavior, internalizing and externalizing behaviors, and conduct disorder, etc.

#### Neurocognition

NIH Cognition Battery Toolbox (*nihtbx*) was used as the measure for general cognitive function (Akshoomoff et al., 2013). It assesses seven cognitive domains, including language vocabulary knowledge, attention, cognitive control, working memory, executive function, episodic memory, and language. Three composite scores, i.e. fluid, crystal, and the total composite scores, were calculated based on domain-specific scores. These 10 scores were available for baseline. Only 6 assessments associated with crystal composite scores were available for FL2.

#### Mental Health

Overall mental health assessments included *Prodromal Psychosis Scale* (pps) (Loewy et al., 2011), *UPPS impulsive behavior scale* (Urgency, Premeditation (lack of), *Perseverance* (lack of), *Sensation Seeking, Positive Urgency, Impulsive Behavior Scale* (Barch et al., 2018), *Behavioral inhibition scale* (BIS) (Carver and White, 1994). These data were available for baseline and FL2.

#### Brain Measures

All children underwent standardized resting-state fMRI, structural MRI, and DTI scans. Acquired images were processed and quality controlled at the *Data Analysis, Informatics and Resource Center* of the ABCD study (Hagler et al., 2019). Network connectivities were calculated as the average fisher-transferred functional connectivity between each pair of ROIs within or between networks (12 networks in total) based on the Gordon atlas (Gordon et al., 2016). In addition, network connectivities between each network and each subcortical region (19 subcortical regions in total (Fischl et al., 2002), see Fig. 3) were also calculated. Cortical area, gray matter volume, and cortical thickness were based on Destrieux Parcellation (148 regions) (Destrieux et al., 2010). Subcortical volume for 36 regions (Fischl et al., 2002) and FA for 35 tracts (Hagler et al., 2009) were calculated. These brain measures were available for baseline and FL2.

### Data Analysis

We first evaluated data from the ABCD dataset based on recommended quality check procedures to ensure data quality. For example, for later resting-state analysis, only participants who passed the rs-fMRI quality check were included for later analysis (n = 9387). Furthermore, we also excluded participants who have missing data for the covariates listed above (n =1064). In the end, among the 11,879 participants, 8323 of them met these criteria. Next, we then perform the propensity score matching procedure (Stuart, 2010) to estimate the causal effects of sleep loss on the outcome measures, implemented through the *MatchIt* R package (Ho et al., 2011). Finally, we performed mediation analyses to investigate how brain changes related to insufficient sleep would mediate the relationship between baseline sleep loss and compromised cognitive and affective functions at different longitudinal timepoints, see Yang, et al., 2021 for detailed processing steps (Yang et al., 2021).

#### Propensity Score Matching

In this procedure, participants were matched based on the probability of being in a comparison group conditioned on observed covariates using logistic regression (Rosenbaum and Rubin, 1985, 1983). We first identified an appropriate propensity model based on both basic demographic characteristics of a participant (e.g., age, sex, the interaction between age and sex, race, location) and theoretically relevant covariates in adolescent sleep patterns (e.g, socioeconomic status such as parent education level and household income, (Felden et al., 2015; Tomasi and Volkow, 2021); puberty status, (Sadeh et al., 2009); BMI, (Meyer et al., 2012)). In addition, we considered the average motion during resting scans (mean framewise-displacement, FD) and the number of fMRI time points that remained after preprocessing as technical covariates, as both head motion and image quality are critical for interpreting pediatric structural and functional neuroimaging (Makowski et al., 2019). Overall, this propensity model captures 11.74% (adjusted R-square) shared variance with sleep sufficiency. Next, we matched participants with sufficient versus insufficient sleep based on one-to-one matching without replacement, in which participants with sufficient sleep (n = 4181) were selected sequentially to be matched with participants with insufficient sleep (n = 4142) from lowest to highest propensity score. We identified these matched samples based on a predefined propensity score radius (i.e., caliper = 0.1) and then compared the standardized mean difference of covariates between these groups of participants to check for the balance (Chan et al., 2016). As demonstrated in the previous research (Austin, 2014), this caliper matching procedure without replacement has good matching performance with less bias in estimating the effects of matched group comparisons that are robust to matching orders and are similar to exact matching (also see Chan et al., 2016 for details). In our current sample, we found that all the covariates were well balanced between comparison groups after this matching (see Supplementary Figure S1), and hence additional group differences could not be attributed to these covariates. In the end, we found 3021 matching pairs based on this procedure, among which 2762 pairs had FL1 data and 749 pairs had FL2 (imaging) data. We then focus our analysis on these matched group comparisons.

#### Group Comparison

For outcome comparison, because matching only reduces between-group variance without making the matched samples truly correlated, independent-sample t-tests were used to examine the matched group difference (SS vs IS groups) on dependent variables, including assessments of problem behaviors, mental health, cognition, and brain measurements of network connectivity, cortical area, gray matter volume, cortical thickness, and structural connectivity (FA). For each brain measurement, independent-sample t-tests were only performed on participants pairs that both passed corresponding quality control for this measurement. False discovery rate (FDR) correction was performed for multiple comparisons for baseline measures. In addition, we chose Cohen’s *d* value 0.15 as an additional threshold for network connectivities and behavior assessments to identify targets for subsequent mediation analysis. This criterion is chosen for various reasons. First, using effect size as a threshold has been shown to improve replicability in neuroimaging findings (Vandekar and Stephens, 2021). Second, a recent study found that effect sizes of association between two variables in ABCD were mostly around 0.03 to 0.09 (Owens et al., 2021). The current criterion of Cohen’s *d =* 0.15 would, therefore, be greater than the common effect size identified in the previous research (Owens et al., 2021).

#### Mediation Analysis

We also performed median analyses to evaluate how the changes in brain structures and functions may be used as a biomarker for the adverse effects of disrupted sleep loss adolescent development identified by matched group comparisons. Here, as sleep duration remains an independent variable, baseline or FL2 behavioral measures identified through matched group comparison were considered outcome variables. We included baseline brain measures as mediators to evaluate whether the structural and functional differences between the two groups (IS and SS) in the brain at baseline can account for later neurocognitive functions after 2 years. Age, sex, race, pubertal status, BMI, parent’s education, household income, data collection sites, mean FD, and the number of time points remaining (for FL2 behvaioral meansures, corresponding baseline behaiviroal measures were added as an additional covariate) were controlled as covariates in the mediation analyses. The significance of these mediation analyses was estimated using bootstrap sampling with 100,000 random-generated samples. We implemented these analyses via the established mediation toolbox (https://github.com/canlab/MediationToolbox) based on previous research (Wager et al., 2009, 2008).

## Results

To investigate the long-lasting effects of insufficient sleep on adolescent neurocognitive development, we identified individuals with sufficient or insufficient sleep based on parental reports of a child’s sleep hours during the initial 6 months before baseline assessment. We matched these two groups based on various key covariates as previously described (e.g., sex, socioeconomic status, etc.; n = 3021 matched pairs, 6042 participants in total) and retained the group assignment for these well-matched pairs in subsequent analyses to allow both cross-sectional and longitudinal comparisons. We examined whether their sleep patterns remain relatively stable throughout the course of the study. As shown in Figure 1, we find that participants’ sleep pattern in the IS group (bottom row) remains relatively stable over the course of 2 years, whereas participants in the SS group (upper row) tend to sleep less and less over time. Because the long-term impact of insufficient sleep may accumulate over time, we expect that participants with initial insufficient sleep show adverse effects on behavioral and brain measures at follow-up time points as compared with the SS group.

**Figure 1.**
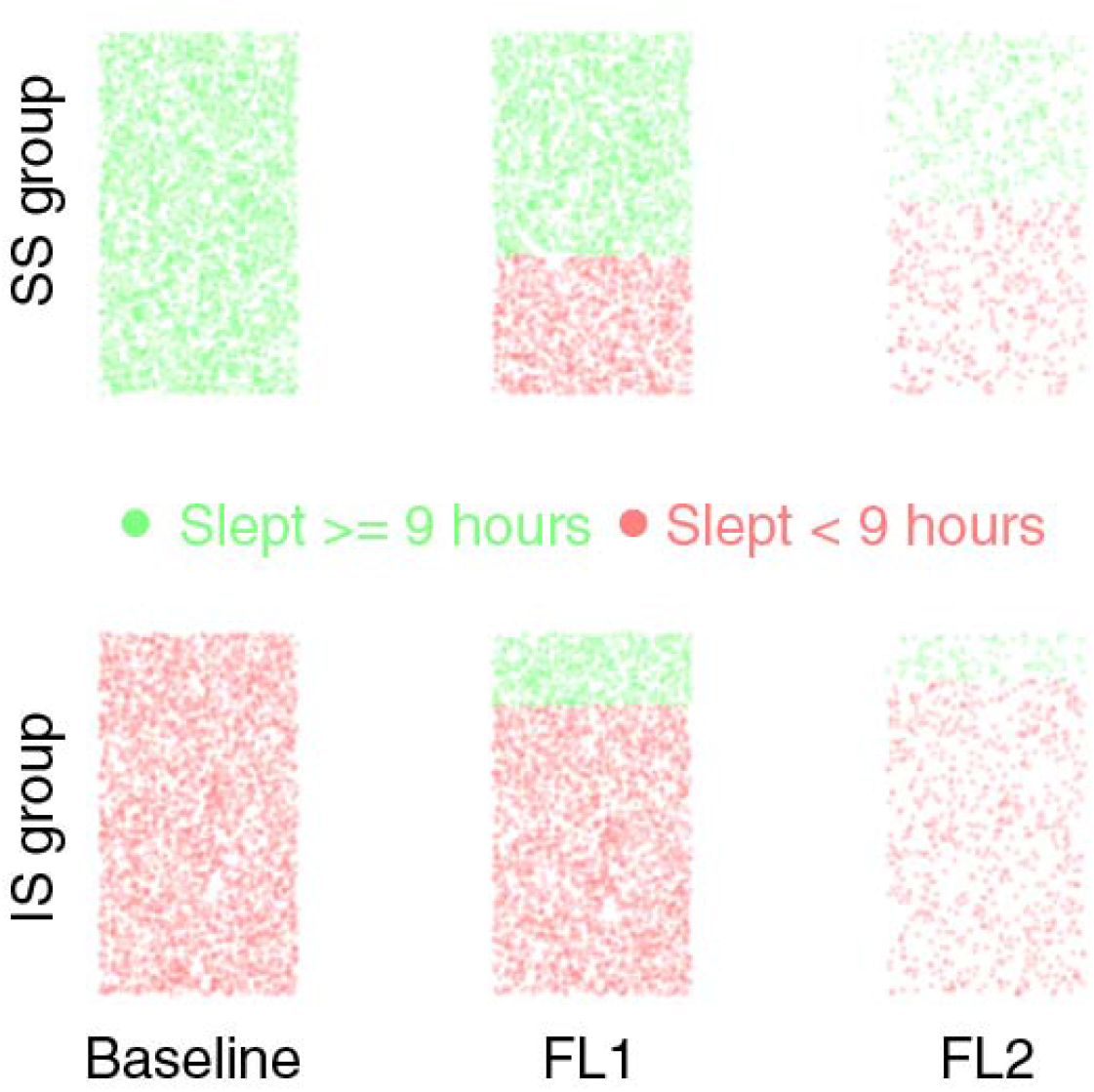
Sleep duration changes in matched SS and IS groups based on baseline assessment. Each dot represents a datapoint for one child. Participants’ sleep pattern in the IS group (bottom row) remains relatively stable over the course of 2 years, whereas participants in the SS group (upper row) tend to sleep less and less over time. Green dot represents a child that slept >= 9 hours per day at the corresponding time point. Red dot represents a child that slept less than 9 hours per day at the corresponding time point. SS: sufficient sleep; IS: insufficient sleep.

### Sleep duration and adolescents’ behaviors, cognition, and mental health over the span of 2 years

We first examined how baseline sleep duration influences the 42 baseline behavioral measurements that capture adolescence’s problematic behaviors (e.g., conduct and psychosis, 20 items), cognitive functions (e.g., fluid and crystal intelligence, 10 items), and mental health (e.g., depression and anxiety, 12 items). Consistent with some previous findings, we found insufficient sleep had widespread impacts on these baseline behavioral measures (Brooks et al., 2021; Cheng et al., 2020). In particular, we found that 32 out of these 42 assessments showed a significant difference between SS and IS groups after propensity score matching on covariates (Figure 2A; *p* < .05, FDR corrected). Among these 32 measures, 4 assessments had an absolute Cohen’s d value (|SS - IS|) higher than an arbitrary cut-off of 0.15, which is greater than 50% of the typical effect sizes identified based on the ABCD dataset (Owens et al., 2021). That is, individuals with sufficient sleep show fewer depression symptoms (cbcl-dsm5-depress, Cohen’s d = -0.20, *p* < .001), fewer psychosis through problems (cbcl-thought problems, Cohen’s d = -0.18, *p* < .001), better picture vocabulary test performance (nihtbx-picture vocabulary test, Cohen’s d = 0.18, *p* < .001), and higher crystal intelligence (nihtbx-crystal cognition, Cohen’s d = 0.17, *p* < .001).

**Figure 2.**
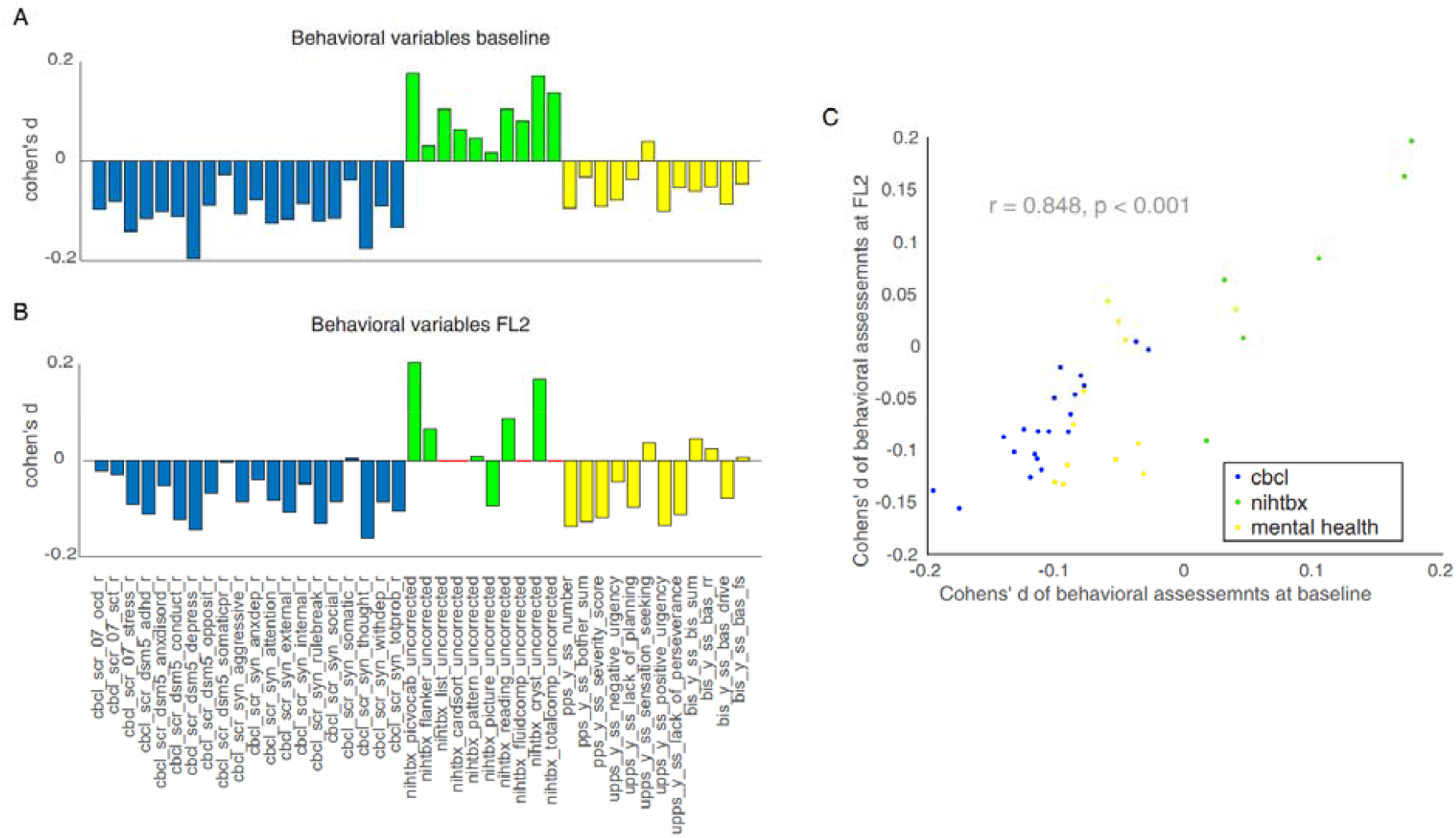
The effect sizes (Cohen’s d) of insufficient sleep on behavioral assessments. Panel A: Baseline behavioral assessments. Positive value means the SS group has higher value than the IS group. Panel B: 2 years follow-up behavioral assessments. Positive value means the SS group has higher value than the IS group. Panel C: Cohen’s d at baseline (x-axis) significantly correlated with Cohen’s d at FL2 (Y axis), *r* = 0.848, *p* < 0.001. Blue: behavioral problems from cbcl; Green: cognition assessments from nihtbx; Yellow: mental health assessments. cbcl: Child Behavior Checklist; nihtbx: NIH Cognition Battery Toolbox. See supplemental table 1 for more information about the variable names. SS: sufficient sleep; IS: insufficient sleep; FL2: 2-year follow-up; red line: 4 measures of cognition were not available for FL2.

We observed similar patterns in the 2-year follow-up, in that 13 measures of the 32 significant behavioral assessments remained statistically significant when the SS and IS groups were compared (*p* < .05, uncorrected post-hoc comparisons), with similar effect sizes as compared with that at baseline comparison. This is supported by a significant correlation between the Cohen’s d of the difference between SS and IS in the 2-year follow-up and that at baseline for all the 42 behavioral measures (Figure 2C; *r* = 0.85, *p* < .001). These results suggest that sleep loss reliably influenced adolescents’ behavior, cognition, and mental health over time. Similar observations are also found in the 1-year follow-up, where only 20 assessments of problem behaviors are available (see Supplementary Figure 2).

### Sleep duration and resting-state functional connectivity in the developing brain

We next examined how sleep duration affects the intrinsic functional organization of brain networks in the developing brain. Out of the 306 unique connections of resting-state functional connectivity, 93 of them show a significant difference between the SS and IS groups (*p* < 0.05, FDR corrected, see Fig. 3A). Among them, 5 connectivity measures have a Cohen’s d value higher than 0.15 (SS - IS), including the auditory network (au) – right putamen (ptrh), Cohens’ d = 0.22, *p* < .001; the cingulo-opercular network (cerc) – left caudate (cdelh), Cohen’s d = 0.19, *p* < .001; the cingulo-parietal network (copa) – right pallidum (plrh), Cohen’s d = 0.16, *p* < .001; the retrosplenial-temporal network (rst) – right cerebellum cortex (crcxrh), Cohen’s d = 0.18, *p* < .001; and the rst – right ventral diencephalon (vtdcrh), Cohen’s d = 0.15, *p* < .001. It is worth mentioning that all of these subcortical regions (except the right cerebellum) are a part of the basal ganglia, which plays a core role in regulating the sleep-wake cycle (Hasegawa et al., 2020). This convergence of disturbed connectivity patterns between cortical functional networks and the basal ganglia in the IS group suggest that the disruption of sleep loss on subcortical brain functions can merge very early, even in the developing brain.

**Figure 3.**
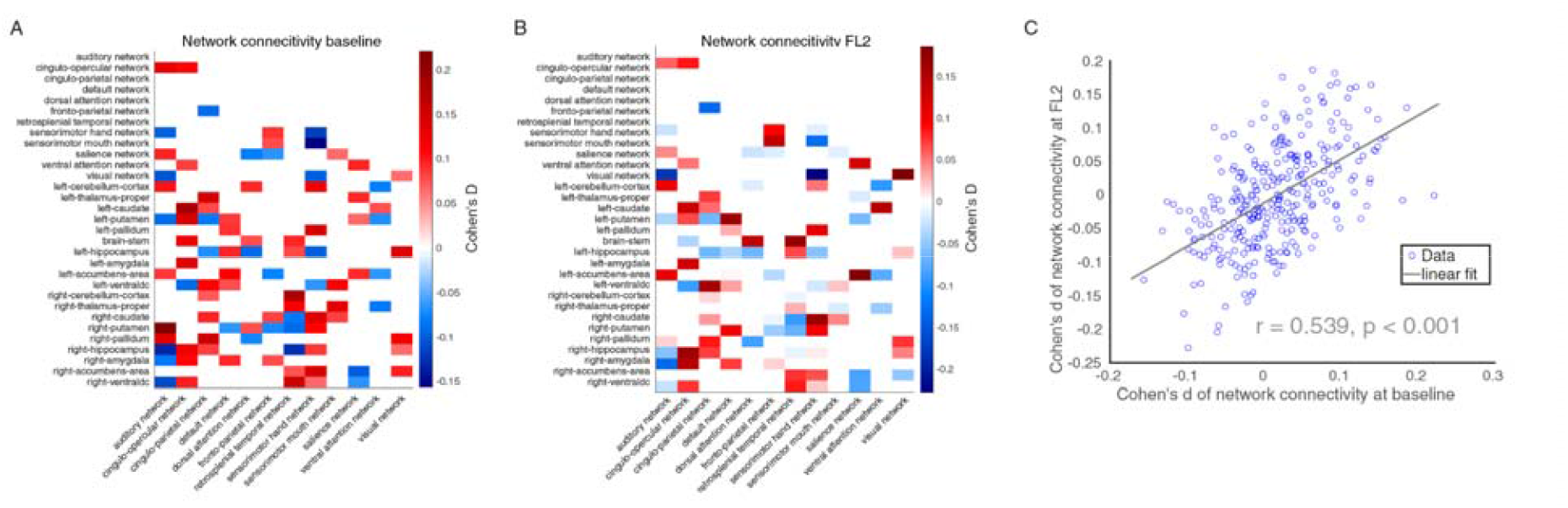
The effect sizes (Cohen’s d) of insufficient sleep on network connectivities. Panel A: Cohen’s d of insufficient sleep on baseline network connectivities, only 93 connections that survived FDR correction shown here. Positive value (red) means the SS group has higher value than the IS group. Negative value (blue) means the SS group has lower value than the IS group. Panel B: Cohen’s d of insufficient sleep on 2-year follow-up network connectivities, 93 connections that survived FDR correction on baseline data shown here. Panel C: Cohen’s d at baseline (x-axis) significantly correlated with Cohen’s d at FL2 (Y axis), *r* = 0.539, *p* < 0.001. All 306 connections were shown in the scatter plot. SS: sufficient sleep; IS: insufficient sleep

To examine whether these findings are robust over time, we analyzed available 2-year follow-up data in the matched pairs. Out of the 93 significant connectivity measures from baseline between-group comparison, 22 of them also reach statistical significance at 2-year follow-up comparison (*p* < .05, uncorrected post-hoc comparisons). Critically, the effect sizes of sleep duration on all 306 available functional connectivity measures are significantly correlated with one another across different points of comparison (i.e., baseline and 2-year follow-up: *r* = 0.54, *p* < .001). These findings suggest that sleep duration reliably influences functional connectivity patterns over the span of 2 years in the developing brain.

### Sleep duration and brain structural changes in the developing brain

We further investigated how sleep duration affects the development of brain structures, in particular, gray matter volume, cortical area, cortical thickness, and FA. For gray matter volume size, 12 out of the 184 anatomical regions parcellated by the ABCD dataset reached statistical significance in the baseline comparison between the SS and IS group (Figure 4; p < 0.05, FDR corrected). However, none of these measures reach a Cohen’s d of 0.15. Yet, we identified 3 regions with Cohen’s d values higher than the typical effect sizes found in the ABCD dataset (0.03 to 0.09; Owens et al., 2021). That is, individuals with sufficient sleep show greater gray matter volume sizes in the bilateral temporal pole in the anterior temporal lobe (Cohen’s d = 0.11 and 0.10, for left/right, respectively, *p*’s < .001) and right anterior cingulate cortex (ACC, Cohen’s d = 0.12, *p* < .001). In 2-year follow-up, we find that these effect sizes are slightly evaluated when SS and IS groups are compared (Cohen’s d for left and right temporal pole = 0.18 and 0.14, respectively, *p*’s < .01; Cohen’s d for ACC = 0.07, *p* = .19). However, longitudinal comparisons did not reveal a significant difference between 2-year follow-up and baseline assessment in the matched pairs that have data at both timepoints (*p*’s > .10). We therefore examine the stability of these effects over time. We find that patterns of Cohen’s d on the gray matter volume in the184 regions at baseline are consistently correlated with that at 2-year follow-up (Figure 4C; *r* = 0.517, *p* < .001), confirming that sleep duration reliably impacts gray volume sizes over time.

**Figure 4.**
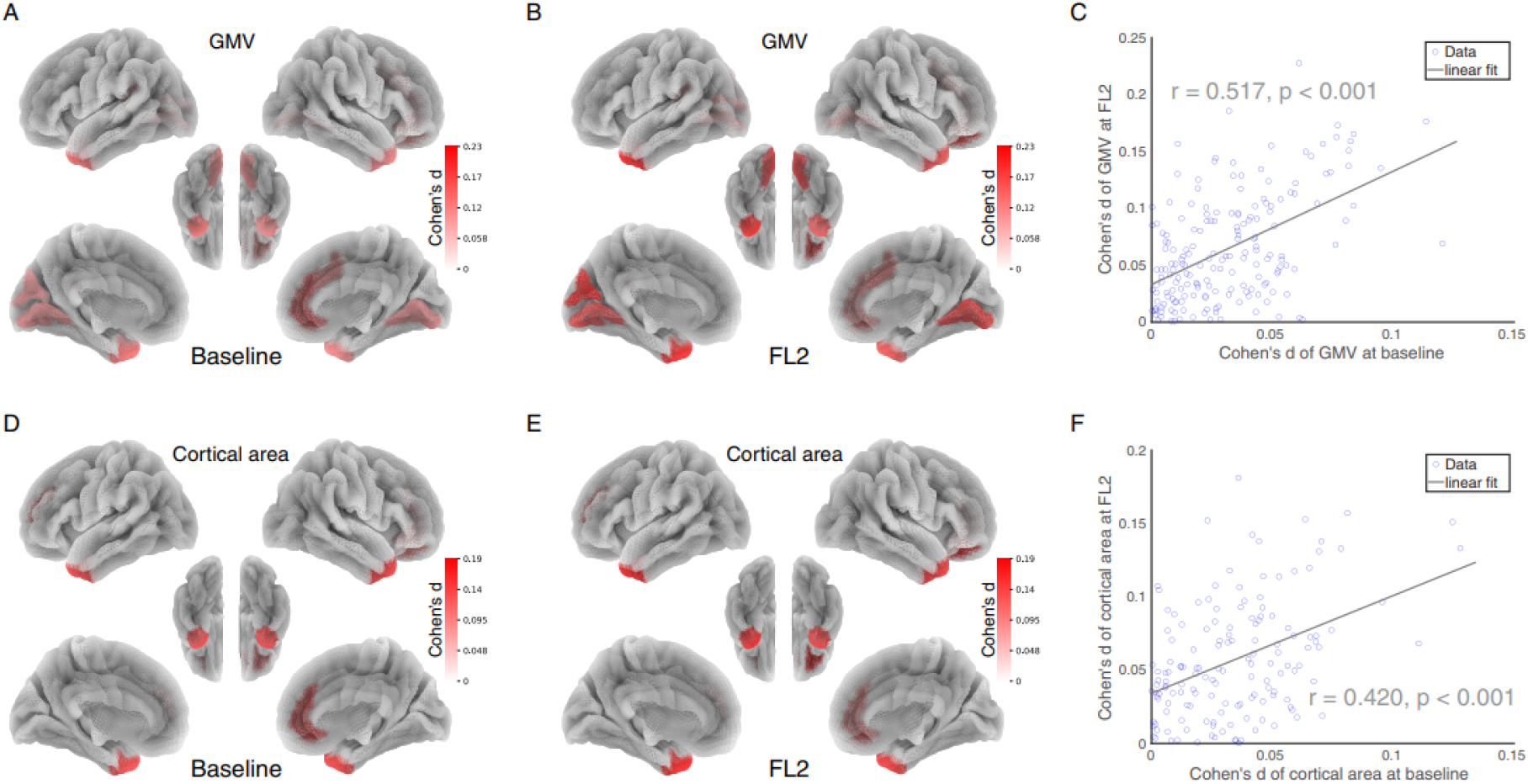
The effect sizes (Cohen’s d) of insufficient sleep on brain structural measurements. Panel A: Cohen’s d of baseline GMV on 12 regions that survived FDR correction. Panel B: Cohen’s d of 2-year follow-up GMV on 12 regions that survived FDR correction on baseline data. Panel C: Cohen’s d of GMV at baseline (x-axis) significantly correlated with Cohen’s d of GMV at FL2 (Y axis), *r* = 0.517, *p* < 0.001. All 186 regions were shown in the scatter plot. Panel D: Cohen’s d of baseline cortical area on 5 regions that survived FDR correction. Panel E: Cohen’s d of 2-year follow-up cortical area on 5 regions that survived FDR correction on baseline data. Panel F: Cohen’s d of cortical area at baseline (x-axis) significantly correlated with Cohen’s d of cortical area at FL2 (Y axis), *r* = 0.420, *p* < 0.001. All 148 regions were shown in the scatter plot. Note: Positive value (red) means the SS group has higher value than the IS group. No negative value was found. Only regions that survived FDR correction were shown in the figure. SS: sufficient sleep; IS: insufficient sleep; GMV: gray matter volume.

The analysis results of sleep effects on cortical area and cortical thickness were consistent with the findings based on gray matter volume size. We found that individuals with sufficient sleep showed larger cortical areas in the bilateral temporal pole (Cohen’s d = 0.13 and 0.13 for left and right, respectively, *p*’s < .001) and right ACC (Cohen’s d = 0.11, *p* < .001) at baseline assessment. Similar findings were observed in 2-year follow up (Cohen’s ds for left and right temporal pole and right ACC are .15, .13, .07, respectively). These observations are also supported by a significant correlation in the patterns of Cohen’s d on the cortical area between baseline and 2-year follow up comparison between SS and IS groups (Figure 4F; *r* = 0.42, *p* < .001). However, for cortical thickness, we did not observe similar patterns in Cohen’s d when SS and IS are compared (See Supplementary Figure 3A; *r* = 0.02, *p* = .81), suggesting that cortical thickness may not be reliable to detect the effects of sleep duration of gray matter changes over time.

FA is a measure of white matter integrity. We did not find statistically significant differences between SS and IS in all 37 regions (all *p* > 0.05, FDR corrected). Yet, the three tracts with the highest cohen’s d values were the left inferior longitudinal fasiculus tract (Cohen’s d = 0.09, *p* < .01, uncorrected), left superior corticostriate tract (Cohen’s d =0.08, *p* < .01 uncorrected), and left superior corticostriate tract-parietal part (Cohen’s d = 0.08, *p* < .01 uncorrected), among which the corticostriate tracts connect the striatum in the basal ganglia with the cerebral cortex. Patterns of Cohen’s d on FA were reliable between baseline and 2-year follow-up comparisons (Supplementary Figure 3B; *r* = 0.571, *p* < .001).

### Changes in resting-state functional connectivity and structural measures mediate the effects of sleep duration on behavioral patterns

Finally, we tested whether the brain measures identified with larger effect sizes in respective categories (i.e. 5 network connectivities, 3 cortical areas, and 3 gray matter volumes) mediate the effects of sleep duration on the 4 behavioral measures identified with a Cohen’s d greater 0.15 (i.e., depression, thought problems, picture-vocabulary test performance, and the composite score of crystal intelligence). As these brain and behavioral measures are identified based on between group comparisons with well-matched covariates using propensity score matching, the effects of sleep duration on each individual measure are unlikely modulate by these covariates (e.g., sex and socioenomic status). Yet, these covariates remain possible to affect that relationship between brain and behavioral variables. Hence, in the mediation analysis, we regressed out these covariances. At baseline, we found that the identified functional and structural brain measures robustly mediated the effects of sleep duration on picture vocabulary test and crystal intelligence (Figure 5A; bootstrap *p* <.05). Three out of five network connectivity measures also mediated the effect of insufficient sleep on depression and thought problems (Fig. 5A; bootstrap *p* < .05). We have also identified distributed mediation patterns across all behavioral measures (see Supplementary Figure S4).

**Figure 5.**
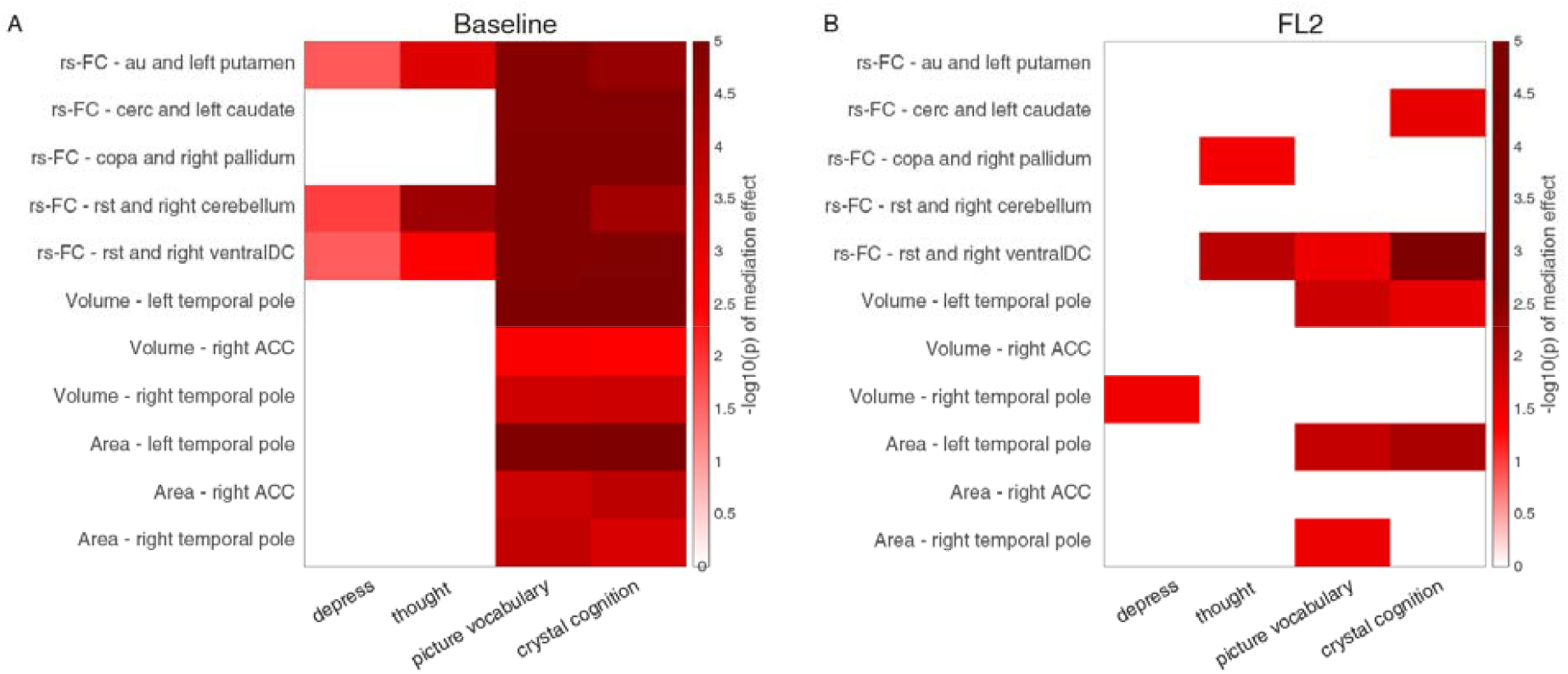
Brain measures mediated the effect of insufficient sleep on behavioral problems and cognition. Panel A: all brain measures mediated the effect of insufficient sleep on picture vocabulary test and crystal cognition at baseline. The log-transferred bootstrapped p-value of the mediation effect were shown. Panel B: brain measures mediated the effect of insufficient sleep on cognition at 2 years follow-up, after controlling for baseline cognition measures. The log-transferred bootstrapped p-value of mediation effect were shown. Only mediation effects reached statistical significance using bootstrap sampling with 100,000 random-generated samples shown in the figure. Note: au: auditory network; cerc: cingulo-opercular network; copa: cingulo-parietal network; rst: retrosplenial-temporal network; ventralDC: ventral diencephalon; ACC: anterior cingulate cortex.

To investigate the robustness of these observations over time, we performed a longitudinal mediation analysis, in which we used baseline brain functional and structural measures to predict the effects of sleep duration on behavioral measures at 2-year follow-up, after partialling covariates and baseline behavioral measures. This time-lag analysis can further reveal whether the identified brain measures can serve as reliable biomarkers of behavioral changes over time. We found that some of the identified functional and structural measures reliably mediated the effects of sleep duration on thought problems (i.e., rst – right ventral diencephalon connectivity), picture vocabulary, and crystal intelligence (i.e., rst – right ventral diencephalon connectivity, left and right temporal pole gray matter measures; see Figure 5 and Supplementary Figure S4).

## Discussion

Here, by controlling for several known factors that covariate with sleep duration, the current study estimated the long-lasting impacts of sleep duration on neurocognitive development in early adolescence. Of primary interest, we identified two important neural mechanisms of sleep loss on neurocognitive development that have not been systematically examined in the literature. First, intrinsic functional connections between the basal ganglia and cortical regions during resting state play an important role in cognitive and affective changes induced by insufficient sleep. Second, structural properties of the temporal pole were involved in the detrimental effect of insufficient sleep on behavioral estimates of crystal intelligence. We found that these effects can last for at least over the span of 2 years, highlighting the long-lasting consequences of insufficient sleep on adolescent neurocognitive development. Our findings, therefore, provide strong empirical and theoretical grounding for early sleep intervention to improve long-term development outcomes in adolescence.

### Sleep duration, cortico-basal ganglia connections, and early adolescence development

The effects of insufficient sleep on network connectivities and white matter connectivities (did not survive FDR correction) converged on the connections between the basal ganglia and cortical regions. Basal ganglia contain a group of subcortical nuclei that regulate core neurotransmitters (e.g., dopamine and adenosine) to support motor control, habit learning, reward processing, and affective processes. Its key role in regulating sleep-wake behavior through cortical activation has also been identified (Lazarus et al., 2013). Hence, it has been hypothesized that sleep loss disrupts the homeostasis of dopamine and adenosine systems, and subsequently impairs the cortex-basal ganglia-thalamus-cortex circuit (Sil’kis, 2014). This can result in weakened attention and limited information processing, leading to impairments to cognitive and affective functions. Here, based on large-scale data with well-matched covariates, we obtain robust neuroimaging evidence supporting this hypothesis. In particular, we identify two core basal ganglia-cortex connections (rs-FC – copa and right pallidum and rs-FC – rst and right ventralDC,) that mediate the impacts of insufficient sleep on depressed mood, thought problems, and crystal intelligence over the span of 1-2 years (see Supplementary Figure S4).

These reliable patterns are in line with recent research efforts in treating resting-state functional connectivity as a trait-like measure to account for individual differences in neurocognitive functions (Finn et al., 2017; Noble et al., 2019).

### Sleep duration, Temporal lobe, and Crystal Intelligence in early adolescence

In contrast to the previously mixed findings on the relationship between sleep loss and higher cognition or intelligence (Astill et al., 2012; Cassoff et al., 2014; de Bruin et al., 2017), we leverage large-scale neural and behavioral data with well-balanced covariates based on propensity scoring matching to reveal the long-lasting impact of sleep duration on crystal intelligence (Cohen’s d = 0.17), which is about twice as compared with that on fluid cognition (Cohen’s d = 0.08). Our mediation and longitudinal mediation analyses show that this behavioral effect is associated with changes in core neural substrates that are related to the representation (temporal pole in the anterior temporal lobe) and retrieval (rst, retrosplenial-temporal network) of structuralized knowledge. As supported by rich prior evidence, the anterior temporal lobe is critical for semantic memory and conceptual knowledge (Bonner and Price, 2013; Herlin et al., 2021; Lambon Ralph et al., 2009; Xie et al., 2020). Temporary disruption of this brain area with transcranial magnetic stimulation can interrupt semantic processing in healthy adults (Lambon Ralph et al., 2009). Consistent with the above evidence, we found that structural properties (i.e. cortical area and gray matter volume) of the temporal lobe mediated the long-lasting detrimental effects of insufficient sleep on picture-vocabulary test and the composite score of crystal cognition. Similarly, the retrosplenial network (rst) that is closely related to the medial temporal lobe system (Kaboodvand et al., 2018) is also affected by sleep loss and associated with the detriment in picture-vocabulary test performance (Figure 5). These findings raise the possibility that sleep loss may compromise memory consolidation processes that involve core temporal lobe structures (e.g., anterior and medial temporal lobe) (Born and Wilhelm, 2012; Tononi and Cirelli, 2014, 2006) in the developing brain. Future research should further articulate how the disturbance of this process under sleep loss would influence the critical development period of memory generalization and the formation of crystal knowledge at even younger ages (Ngo et al., 2021).

### Estimating the impacts of sleep on neurocognitive development

Another important contribution of this study is to estimate the causal impacts of sleep loss on neurocognitive development in early adolescence based on propensity matching that mimics randomized experiments based on observational data. As core covariates that may influence sleep duration and neurocognitive measures are well balanced between SS and IS groups (e.g., household income), the group comparison after matching therefore could not be confounded by these covariates. This analytical approach, therefore, resolves the uncertainty in the literature on whether the impacts of sleep duration on neurocognitive development would be confounded by socioeconomic variables like household income (Dutil et al., 2018). As shown in Supplementary Figure S1, this covariate indeed can significantly predict whether a child would have sufficient or insufficient sleep, such that its significant covariance on neurocognitive development outcomes may not be adequately handled by the regular regression model. By incorporating propensity matching as recommended by the ABCD study (Dick et al., 2021), our study clarify the effects of sleep loss on neurocognitive development that is void of the confounds from parent education level and household income (Felden et al., 2015; Tomasi and Volkow, 2021), puberty status (Sadeh et al., 2009), BMI (Meyer et al., 2012), sex (Lee et al., 1999), motion during resting-state fMRI scan (Makowski et al., 2019).

### Caveats and future directions

Despite the strength in matching for various observed covariates, the propensity score matching approach could not fully control for unobserved covariates that may influence adolescents’ sleep patterns and neurocognitive development. For example, in the current ABCD dataset, measures concerning a child’s school programs are limited. As different school districts may implement different programs to improve students’ sleep habits, our results may be influenced by these unaccounted for covariates. To fully adjust for these covariates, future studies may examine how sleep manipulation influences neurocognitive in early adolescence based on a longitudinal design. For example, it is of great translational interest to test whether sleep extension can reverse the effect of insufficient sleep on behavior and brain measurements identified in this study. Last, our current brain measures (i.e., resting-state functional connectivity and structure measures) are task-free, which may not capture some information related to specific abilities (e.g., memory consolidation involving the medial temporal lobe system). Future research can further examine how sleep duration moderate the task-based brain and behavioral measures in the ABCD dataset.

## Conclusion

By applying propensity score matching to the large-scale ABCD dataset, this study estimates the long-lasting impacts of sleep loss on neurocognitive development in early adolescence while controlling key covariates, including sex, socioeconocmic status, purberty status, and physical health indicator (i.e., BMI), etc. Our analyses suggest that (1) cortico-basal ganglia connections might play an important role on the board behavioral effects of sleep loss on cognitive and affective functions and (2) structure properties of the anterior temporal lobe might contribute to the effect of sleep loss on crystal cognition. These effects can last at least over the span of 2 years, highlighting the importance of early sleep intervention at young ages to improve long-term neurocognitive development outcomes in adolescence.

## Data Availability

The ABCD data that are used by this study are available in National Institutes of Mental Health Data Archive (NDA): https://nda.nih.gov/abcd.

## Author contributions

F.N.Y. conceptualized the study, analyzed the data, generated figures and wrote the original draft. W.X. contributed to conceptualization and visualization, and edited the manuscript. Z.W. supervised study and interpretation, and edited the manuscript.

## Ethics approval statement

ABCD study received ethical approval in accordance with the ethical standards of the 1964 Declaration of Helsinki.

## Acknowledgment

Research efforts in this work were supported by NIH grants: R01AG060054, R01 AG070227, R01EB031080-01A1, P41EB029460-01A1. We thank the ABCD consortium and NIH for providing the data for performing the research in this work. Data used in the preparation of this article were obtained from the ABCD Study (https://abcdstudy.org/) and are held in the NIMH Data Archive. This is a multisite, longitudinal study designed to recruit more than 10,000 children aged 9–10 and follow them over 10 years into early adulthood. The ABCD Study is supported by the National Institutes of Health (NIH) and additional federal partners under award numbers U01DA041022, U01DA041028, U01DA041048, U01DA041089, U01DA041106, U01DA041117, U01DA041120, U01DA041134, U01DA041148, U01DA041156, U01DA041174, U24DA041123, and U24DA041147. A full list of supporters is available at https://abcdstudy.org/federal-partners/. A listing of participating sites and a complete listing of the study investigators can be found at https://abcdstudy.org/principal-investigators/. ABCD consortium investigators designed and implemented the study and/or provided data but did not necessarily participate in analysis or writing of this report. This manuscript reflects the views of the authors and may not reflect the opinions or views of the NIH or ABCD consortium investigators.

